# Global Burden, Risk Factors, Causative Organisms and Antibiotic Susceptibility Patterns in Bacterial Keratitis

**DOI:** 10.1101/2025.09.15.25335752

**Authors:** Rohan Bir Singh, Uday Pratap Singh Parmar, Fasika Woreta, Diyva Srikumaran, Namrata Sharma, Vishal Jhanji

## Abstract

**Background:** Bacterial keratitis (BK) is a leading cause of corneal ulceration and vision loss worldwide. We aimed to synthesize global data on BK prevalence, risk factors, causative pathogens, and antibiotic susceptibility through a systematic review and meta-analysis.

**Methods:** This review (PROSPERO CRD420251088592) searched PubMed, Scopus, Cochrane CENTRAL, and PsycINFO through July 1, 2022, for observational studies of microbiologically confirmed infectious keratitis. Eligible studies reported numbers of BK cases and bacterial antibiotic susceptibility. Two reviewers screened studies and extracted data. The primary outcome was the pooled proportion of keratitis cases attributable to bacteria (random-effects meta-analysis); reported risk factors and susceptibilities were summarized descriptively.

**Results:** Of 5,180 records screened, 58 studies (>127,000 patients) met inclusion criteria. The pooled BK prevalence was 44% (95% CI 39–49%) with high heterogeneity (I²≈97%); prevalence varied by region (e.g. ∼61% in Oceania vs ∼29% in Africa). Among ∼25,000 bacterial isolates, 72% were Gram-positive. The most common species were *Staphylococcus epidermidis* (∼15%), *Streptococcus pneumoniae* (∼13%), *Staphylococcus aureus* (∼10%), and *Pseudomonas aeruginosa* (∼17%). Reported predisposing factors included trauma (∼44% of cases) and contact lens wear (∼23%). Gram-positive bacteria were >95% susceptible to vancomycin; Gram-negatives were >90% susceptible to aminoglycosides and cephalosporins. Fluoroquinolone susceptibility was lower (e.g. ciprofloxacin ∼77% for *S. aureus*); about 20% of *S. aureus* isolates were MRSA.

**Conclusions:** Approximately half of infectious keratitis cases worldwide are bacterial, though the burden varies by setting. The findings emphasize trauma and lens hygiene as key targets for prevention. The predominance of a few organisms and rising resistance patterns underscores the need for broad empirical therapy (covering MRSA and *Pseudomonas*). Strengths include the large multinational dataset; limitations include high between-study heterogeneity and sparse data from some regions. These results can inform clinical management and BK prevention strategies.

**Funding:** No funding was received for this study.

**Research in context:** *Evidence before this study:* We searched PubMed/MEDLINE, Scopus, Cochrane CENTRAL, and APA PsycINFO from January 1, 1975, to July 1, 2022, using combinations of the terms “infectious keratitis,” “microbial keratitis,” “corneal ulcer,” and “bacterial keratitis.” We screened titles, abstracts, and full-text articles for studies reporting microbiologically confirmed cases of microbial keratitis with extractable data on bacterial etiology or antibiotic susceptibility. Only English-language human studies were included; case reports, animal studies, and editorials were excluded. Additional studies were identified by manually reviewing reference lists from eligible articles and relevant reviews. Risk of bias was assessed using an adapted GRADE scoring system for prevalence studies. Prior to this study, most available evidence came from single-center studies or regional case series, and there was no comprehensive global synthesis of the prevalence, microbiological profiles, or antimicrobial resistance patterns in bacterial keratitis. No prior systematic review has quantified the global pooled prevalence of bacterial keratitis across multiple continents or assessed the variation in risk factors and susceptibility patterns.

*Added value of this study:* This is the first global systematic review and meta-analysis to synthesize data on the prevalence, risk factors, causative organisms, and antibiotic susceptibility profiles of bacterial keratitis. We analyzed over 127,000 cases of microbial keratitis from 58 studies across 32 countries, including more than 25,000 bacterial isolates and 5,000 antimicrobial susceptibility profiles. Our findings provide reliable pooled estimates of bacterial keratitis prevalence globally (44%), delineate dominant bacterial species (such as *S. epidermidis*, *P. aeruginosa*, *S. pneumoniae*), and identify leading modifiable risk factors (such as trauma, contact lens wear). We also report concerning trends in antimicrobial resistance, including rising methicillin resistance and declining fluoroquinolone susceptibility.

*Implications of all the available evidence:* These findings highlight bacterial keratitis as a global public health concern, particularly in regions with high trauma or contact lens use. The wide variability in prevalence and species distribution underscores the need for regionally tailored prevention and treatment strategies. The declining efficacy of fluoroquinolones and increasing multidrug resistance among ocular pathogens call for urgent attention to antimicrobial stewardship and the development of updated, evidence-based empirical treatment protocols. Our results emphasize the importance of strengthening global surveillance efforts and enhancing microbiological diagnostic capacity, especially in under-resourced settings.

## Introduction

Corneal opacity is the fifth leading cause of blindness globally.^1^ Infectious keratitis (IK) is a major contributor of corneal blindness, causing an estimated 1.5–2.0 million new cases of monocular blindness annually.^2–5^ According to WHO, approximately 6 million people suffer moderate-to-severe vision impairment from corneal scarring, much of which is attributable to prior infections caused by bacterial, fungal, viral, or parasitic organisms.^6,7^ Bacterial keratitis (BK) is the most common form of IK and patients typically present with pain, redness, and corneal ulceration, and can rapidly cause vision loss if not promptly treated.^8^

The epidemiology of BK varies by geography and local risk factors. Major risk factors include contact lens wear, ocular trauma (especially vegetative injury), ocular surface disease (such as dry eye, blepharitis), eyelid malposition, and prior surgery.^8,9^ In developed countries, contact lens–related BK (often caused by *Pseudomonas* spp.,) the most common underlying risk factor, whereas in tropical or agricultural settings, trauma-related infections (caused by Gram-positive bacteria like Streptococci and Staphylococci) predominate. Gram-positive bacteria are more frequently recovered in BK isolates in temperate regions, while Gram-negative bacteria (and fungi) are relatively more common in tropical climates. Despite these known patterns, global data on BK burden are fragmented. Prompt antibiotic treatment can often limit the loss of vision caused by BK, but rising antimicrobial resistance in ocular isolates is a cause of increasing concern. Globally, there is an increase in the reporting of methicillin-resistant *S. aureus* (MRSA) and fluoroquinolone-non-susceptible *Staphylococcus* or *Pseudomonas*, limiting the current therapies for effectively treating BK.^10–14^ Existing reports of ocular isolates suggest evolving resistance; however, the evidence is limited to single-center or regional distribution of cases. Thus, the global burden of BK and the current antibiotic susceptibility patterns across the world remain unclear.

This systematic review and meta-analysis were performed to assess the global patterns of incidence, risk factors and susceptibility in BK patients. Our objectives were to estimate the pooled proportion of microbial keratitis cases due to bacteria, map geographic variations, identify the risk factors causing BK and predominant causative bacteria, and summarize their antibiotic susceptibility patterns. The findings will inform clinicians and public health authorities about the global impact of BK and guide empirical therapy decisions.

## Methods

This study is a systematic review and meta-analysis conducted in accordance with a pre-registered protocol (PROSPERO registration number: CRD420251088592) and reported following the PRISMA 2020 guidelines. We aimed to synthesize global data on the prevalence and microbiological patterns of BK, as well as the in vitro antibiotic susceptibility of bacterial isolates implicated in IK.

We systematically searched four electronic databases, PubMed/MEDLINE, Scopus, Cochrane CENTRAL, and APA PsycINFO, for studies published from January 1, 1975, through July 1, 2022. **(Figure 1)** The search strategy included the following terms and their synonyms: “infectious keratitis,” “microbial keratitis,” “corneal ulcer,” and “bacterial keratitis.” No restrictions were applied with respect to geography or publication date. The full search strategies for each database are provided in the **Appendix S1**. Additional studies were identified by manually screening the reference lists of included articles and relevant reviews. Only articles published in English and involving human subjects were considered. Grey literature, unpublished datasets, unpublished data and non-English publications were excluded.

**Figure 1.**
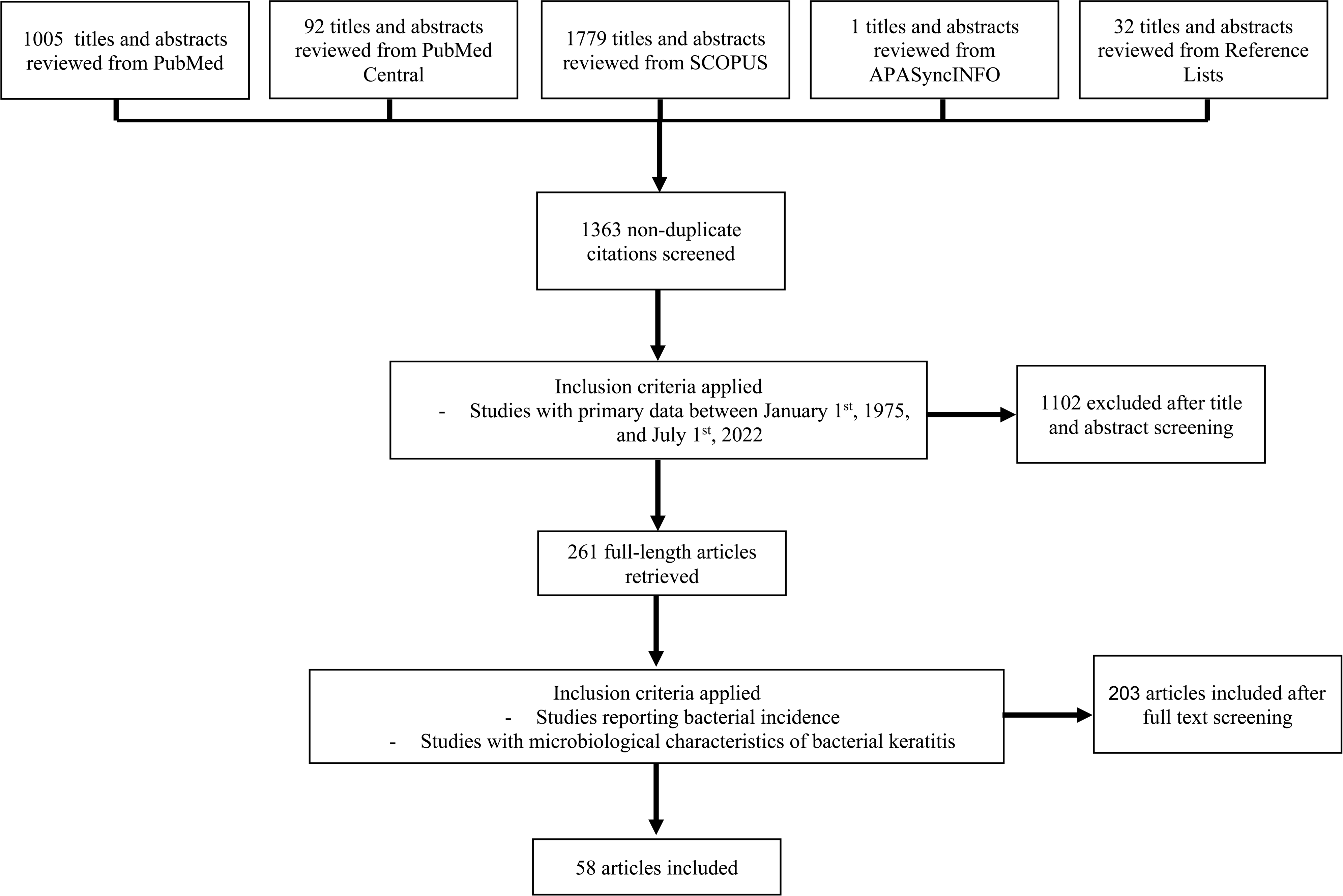
PRISMA 2020 Flow Diagram for Study Selection. Flow of information through the phases of the systematic review. A total of 1363 non-duplicate records were screened after database and reference list searches (PubMed, Scopus, Cochrane CENTRAL, APA PsycINFO). Following title/abstract screening and full-text eligibility assessment, 205 articles were included in the final qualitative and quantitative synthesis based on predefined inclusion criteria: (1) primary data on infectious keratitis between January 1, 1975, and July 1, 2022; (2) reports of bacterial incidence or microbiological characteristics of bacterial keratitis.

We included observational studies, specifically prospective or retrospective cohort studies, case series, and cross-sectional studies, that reported microbiologically confirmed cases of infectious keratitis. To be included in the prevalence meta-analysis, a study had to report both the total number of microbial keratitis cases and the number or proportion attributable to bacteria. For the analysis of antibiotic susceptibility, we included studies that provided in vitro susceptibility data for bacterial isolates obtained from keratitis cases. Case reports, reviews, editorials, animal studies, and studies lacking extractable numeric data were excluded.

Two independent reviewers screened titles and abstracts, assessed full-text eligibility, and extracted data into a standardized form. Discrepancies were resolved by consensus or through adjudication by a third reviewer. Extracted data included study location, time period, clinical setting, study design, and characteristics of the patient population. We also recorded the total number of keratitis cases, the number and proportion of cases caused by bacteria, the identity of bacterial isolates (including Gram stain and genus/species when available), and antibiotic susceptibility profiles (reported as percent susceptible by antimicrobial agent). In studies reporting multiple isolates per patient, each isolate was counted separately. When duplicate data from overlapping populations were identified, the most recent or comprehensive dataset was retained. All data used were aggregated summary-level estimates; no individual patient-level data were sought.

The quality of included studies was assessed using an adapted GRADE (Grading of Recommendations Assessment, Development and Evaluation) framework tailored for prevalence studies. **(Appendix S2)** Key domains evaluated included diagnostic accuracy of microbial identification methods, adequacy of patient sample size, recency of data collection, and clarity in reporting the proportion of BK among all IK cases.^15^ Sensitivity analyses were conducted to explore the impact of excluding studies at high risk of bias.

Our primary outcome was the pooled proportion of microbial keratitis cases caused by bacteria. Meta-analyses were performed using R statistical software (version 4.1.2, R Foundation for Statistical Computing, Vienna, Austria) and the ‘meta’ package. To stabilize variance in proportions, we applied the Freeman–Tukey double-arcsine transformation. Pooled prevalence estimates were calculated using a DerSimonian–Laird random-effects model due to anticipated heterogeneity between studies. Estimates were back transformed to proportions for interpretation. Statistical heterogeneity was assessed using the I² statistic, with thresholds of 25%, 50%, and 75% representing low, moderate, and high heterogeneity, respectively. Where sufficient data were available, subgroup analyses were conducted by geographic region and clinical setting.

As part of our systematic review, we extracted data on predisposing risk factors for BK from studies that reported patient-level or group-level data on relevant exposures. Eligible studies included those that identified specific risk factors such as trauma, contact lens wear, prior ocular surgery, ocular surface disorders, and systemic or local comorbidities in their patient cohorts. For inclusion in the risk factor sub-analysis, studies were required to report either absolute numbers or proportions of BK cases associated with each risk factor. Risk factor data were aggregated across studies, and pooled percentages were calculated as the proportion of total cases reporting each risk factor out of the total number of risk factor-specific events observed. Because risk factor data were inconsistently reported across studies, a formal meta-analysis was not performed; rather, findings are presented descriptively, and Pearson’s correlation analysis was performed.

Due to methodological heterogeneity in antimicrobial susceptibility testing, including differences in laboratory techniques and reporting standards, we did not perform a meta-analysis of antibiotic resistance data. Instead, we summarized the range of reported susceptibilities by agent and organism and developed a global heatmap categorizing antimicrobial activity as high (≥90% susceptible), moderate (50–89%), or low (<50%). There was no funding source for this study, and there was no role of an any other individual or entity, except the authors in study design, data collection, data analysis, data interpretation, or writing of the report.

## Results

We identified 5,180 unique records through our database search. After title and abstract screening, 5,000 records were excluded, leaving 180 full-text articles assessed for eligibility. Among these, 122 studies were excluded primarily due to lack of extractable numerical data (n=58), ineligible study design (n=34), or duplication (n=30). A total of 58 studies were included in the systematic review and meta-analysis of BK prevalence.

The included studies were published between 1975 and 2022 and represented 32 countries across six continents. The largest number of studies came from Asia (n=22), followed by the Americas (n=11), Europe (n=9), Oceania (n=6), and Africa (n=3). **(Table 1)** Most studies were conducted in tertiary ophthalmic centers, though a few were population-based. The sample sizes in each of the studies varied widely, ranging from 20 to 12,000 cases of microbial keratitis per study, with a median of approximately 400 BK cases per study. Collectively, these studies encompassed over 127,000 patients with infectious keratitis, from which 58 provided extractable data on bacterial etiology (n ≈ 56,000). Among these, 54 studies provided species-level bacterial identification (n=25,209 isolates), and 27 studies reported antibiotic susceptibility data (∼5,000 isolates).

**Table 1.** Country- and region-level prevalence of bacterial keratitis among patients with microbial keratitis: pooled estimates from 58 studies (1979–2022).

| Region | Cases | Sample Size | Pooled Prevalence | 95% CI |
| --- | --- | --- | --- | --- |
| **Africa** | 55 | 247 | 22.27 | 17.53 - 27.86 |
| Egypt | 55 | 247 | 22.27 | 17.53 - 27.86 |
| **Asia** |  |  |  |  |
| China | 3011 | 11259 | 26.74 | 25.93 – 27.57 |
| India | 20617 | 74477 | 27.68 | 26.36 – 28.03 |
| Iraq | 162 | 396 | 40.91 | 36.18 – 45.82 |
| Israel | 415 | 943 | 44.01 | 40.87 - 47.19 |
| Japan | 239 | 424 | 56.36 | 51.61 – 61.01 |
| Nepal | 256 | 405 | 63.21 | 58.51 - 67.91 |
| Oman | 182 | 606 | 30.03 | 26.38 - 33.68 |
| Philippines | 185 | 348 | 53.16 | 47.92 - 58.40 |
| Saudi Arabia | 277 | 1062 | 26.08 | 23.53 – 28.80 |
| Singapore | 224 | 531 | 42.18 | 37.98 - 46.39 |
| South Korea | 208 | 379 | 54.88 | 49.87 - 59.89 |
| Taiwan | 1375 | 2488 | 55.26 | 53.30 – 57.20 |
| Thailand | 150 | 280 | 53.57 | 47.73 - 59.41 |
| Turkey | 175 | 620 | 28.23 | 24.68 - 31.77 |
| **Europe** |  |  |  |  |
| Germany | 1162 | 6468 | 17.97 | 17.03 - 18.90 |
| United Kingdom | 2855 | 7457 | 38.28 | 37.18-39.39 |
| **North America** |  |  |  |  |
| Canada | 2265 | 4312 | 52.53 | 51.04 - 54.02 |
| Mexico | 544 | 1638 | 33.21 | 30.93 - 35.49 |
| United States of america | 2320 | 4471 | 51.89 | 50.43 - 53.35 |
| **Oceania** |  |  |  |  |
| Australia | 684 | 1052 | 65.01 | 62.08 – 67.84 |
| New Zealand | 249 | 368 | 67.66 | 62.72 – 72.23 |
| **South America** |  |  |  |  |
| Brazil | 2699 | 6804 | 39.67 | 38.51 - 40.83 |
| Paraguay | 267 | 660 | 40.45 | 36.71 - 44.20 |

Across all studies, the proportion of microbial keratitis cases identified as bacterial ranged from approximately 10% to 85%. **(Table 1)** The pooled estimate of BK prevalence was 44% (95% CI 39–49%, p < 0.0001), as shown in the forest plot. Between-study heterogeneity was high (I² = 97.4%), reflecting substantial variability across geographic regions and clinical settings. **(Figure 2)** Geographic variation was pronounced: studies from Oceania reported the highest pooled prevalence of BK (61%; 95% CI 55–66%), with particularly high proportions observed in New Zealand (67.7%; 95% CI 62.7–72.2%) and Australia (65.0%; 95% CI 62.1–67.8%). In contrast, the pooled prevalence in Europe and Africa was lower, at 32% (95% CI 25–39%) and 29% (95% CI 19–39%), respectively. **(Figure 3)** North America and Asia showed intermediate rates, with pooled prevalence estimates of 44% (95% CI 35–52%) and 51% (95% CI 45–57%), respectively. These findings suggest that on average, approximately half of microbial keratitis cases worldwide are attributable to bacteria, but regional “hotspots” and “coldspots” exist, likely reflecting variations in local risk factors such as contact lens usage and antimicrobial practices.

**Figure 2.**
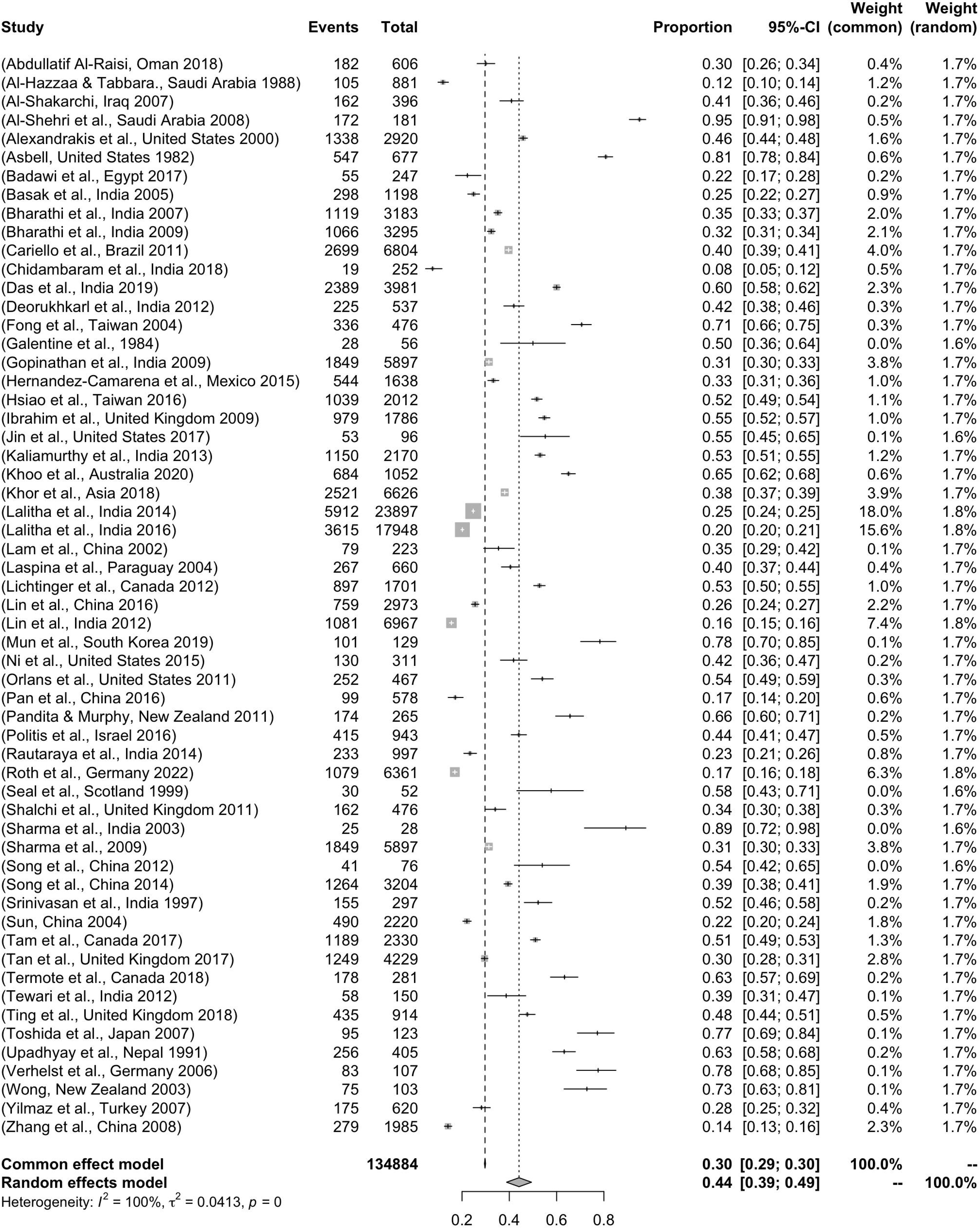
Forest Plot of Proportion of Bacterial Keratitis Among Microbial Keratitis Cases. Proportions with 95% confidence intervals (CIs) for the prevalence of bacterial keratitis among microbial keratitis cases across 58 included studies. The pooled prevalence was 44% (95% CI: 39–49%) using a DerSimonian–Laird random-effects model. Substantial heterogeneity was observed across studies (I² = 100%, τ² = 0.0413, p < 0.001). Study weights under both common-effect and random-effects models are displayed.

**Figure 3.**
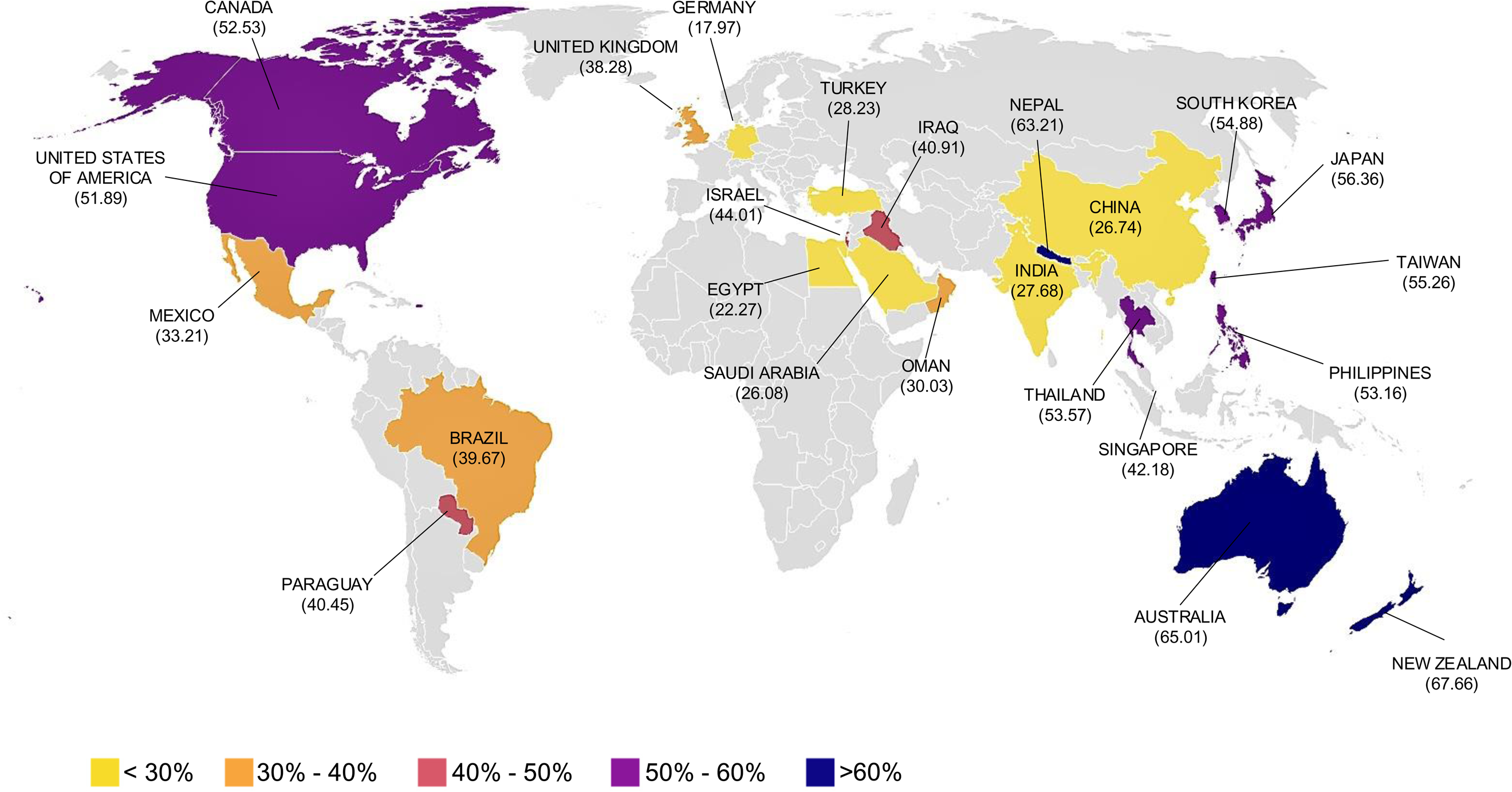
Global distribution map illustrating the pooled proportion of infectious keratitis cases attributable to bacteria by country, based on included studies. Countries are color-coded by prevalence intervals: <30%, 30–40%, 40–50%, 50–60%, and >60%. Notable variation is observed geographically, with the highest prevalence estimates reported from Australia (65.01%), New Zealand (67.66%), and Nepal (63.21%).

Pathogen data from 54 studies (25,209 BK isolates) indicated that Gram-positive cocci predominated (72% of isolates), followed by Gram-negative rods (27%) and Gram-positive bacilli (4%). **(Table 2)** The most isolated genera were *Staphylococcus*, *Pseudomonas*, *Streptococcus*, *Corynebacterium*, and Moraxella. **(Appendix S3)** *Staphylococcus epidermidis* was the most frequently identified species, comprising 15–16% of all isolates, followed by *Streptococcus pneumoniae* (∼13%) and *Staphylococcus aureus* (∼10%). These three species alone accounted for ∼39% of all isolates. *Pseudomonas aeruginosa* was the most common Gram-negative organism (∼17%) and was particularly prevalent in tropical regions and among contact lens users. Together, the four dominant species: *P. aeruginosa*, *S. epidermidis*, *S. pneumoniae*, and *S. aureus*— accounted for over half of all BK isolates. Regional variations were also observed in species prevalence; *P. aeruginosa* was most common in Malaysia, Iran, and Taiwan, whereas *coagulase-negative Staphylococcus* was predominant in the UK and Australia.

**Table 2.** Distribution and prevalence of bacterial organisms isolated from keratitis cases by Gram staining classification.

| Bacterial Species and Classification | Cases | Percentage Prevalence | 95% CI |
| --- | --- | --- | --- |
| **Gram Negative Bacilli** |  |  |  |
| Acinetobacter baumannii | 7 | 0.0169 | 0.01 - 0.04 |
| Enterobactor spp | 12 | 0.0285 | 0.02 - 0.05 |
| Hemophilus influenzae | 23 | 0.0559 | 0.04 - 0.08 |
| Klebsiella oxytoca | 8 | 0.0202 | 0.01 - 0.04 |
| Proteus mirabilis | 2 | 0.0049 | 0.00 - 0.02 |
| Pseudomonas aeruginosa | 6912 | 17.030 | 16.67 - 17.40 |
| Serratia marcescens | 211 | 0.5200 | 0.45 - 0.59 |
| **Gram Negative Cocci** |  |  |  |
| Moraxella spp | 854 | 2.1045 | 1.97 - 2.25 |
| **Gram Positive Bacilli** |  |  |  |
| Corynebacterium spp. | 587 | 1.4453 | 1.33 - 1.57 |
| Nocardia spp. | 279 | 0.6862 | 0.61 - 0.77 |
| **Gram Positive Cocci** |  |  |  |
| Miccrococcus spp. | 148 | 0.3647 | 0.31 - 0.43 |
| Staphylococcus aureus | 4083 | 10.058 | 9.77 - 10.36 |
| Staphylococcus epidermidis | 6271 | 15.450 | 15.10 - 15.81 |
| Staphylococcus simulans | 42 | 0.1032 | 0.08 - 0.14 |
| Streptococcus pneumoniae | 5193 | 12.795 | 12.47 - 13.12 |
| Streptococcus pyogenes | 338 | 0.8339 | 0.75 - 0.93 |
| Streptococcus viridans | 239 | 0.5888 | 0.52 - 0.67 |

Risk factor data were reported in studies encompassing 23,041 patients with bacterial keratitis for whom predisposing conditions were identified. **(Table 3)** Trauma emerged as the most frequently reported risk factor, accounting for 10,179 cases (44.2%). Contact lens wear was the second most common, implicated in 5,233 cases (22.7%), followed by prior ocular surgery in 3,487 cases (15.1%). Other less common predisposing factors included ocular surface disorders (1,844 cases, 8.0%), dry eye disease (604 cases, 2.6%), trichiasis (344 cases, 1.5%), and eroding sutures (291 cases, 1.3%). Systemic comorbidities such as diabetes mellitus were reported in 279 cases (1.2%), and the use of postoperative soft contact lenses was noted in 264 cases (1.2%). Additional factors included blepharitis (226 cases, 1.0%), previous keratitis (183 cases, 0.8%), trachoma (80 cases, 0.35%), herpes simplex infection (15 cases, 0.06%), and glaucoma (12 cases, 0.05%). Among studies reporting correlation coefficients between specific risk factors and the likelihood of bacterial keratitis, trauma showed a moderate inverse correlation with BK (r = –0.2962), suggesting that traumatic etiologies may be more commonly associated with non-bacterial keratitis in certain regions. In contrast, contact lens wear demonstrated a positive correlation (r = +0.1182), consistent with the high frequency of *Pseudomonas aeruginosa* infections observed among contact lens users. Prior ocular surgery was associated with a stronger inverse correlation (r = –0.4485), which may reflect the protective effect of perioperative antibiotic prophylaxis or the higher prevalence of fungal and polymicrobial keratitis in postoperative settings. Notably, we observed that BK caused by improper contact lens use was more common in higher income countries, whereas ocular trauma associated BK was more common in lower income countries **(Figure 4)** These findings highlight substantial variability in predisposing risk factors for BK across geographic and clinical settings. While trauma and contact lens wear remain the leading contributors globally, their relative importance may differ based on environmental exposures, healthcare access, and regional pathogen profiles.

**Figure 4.**
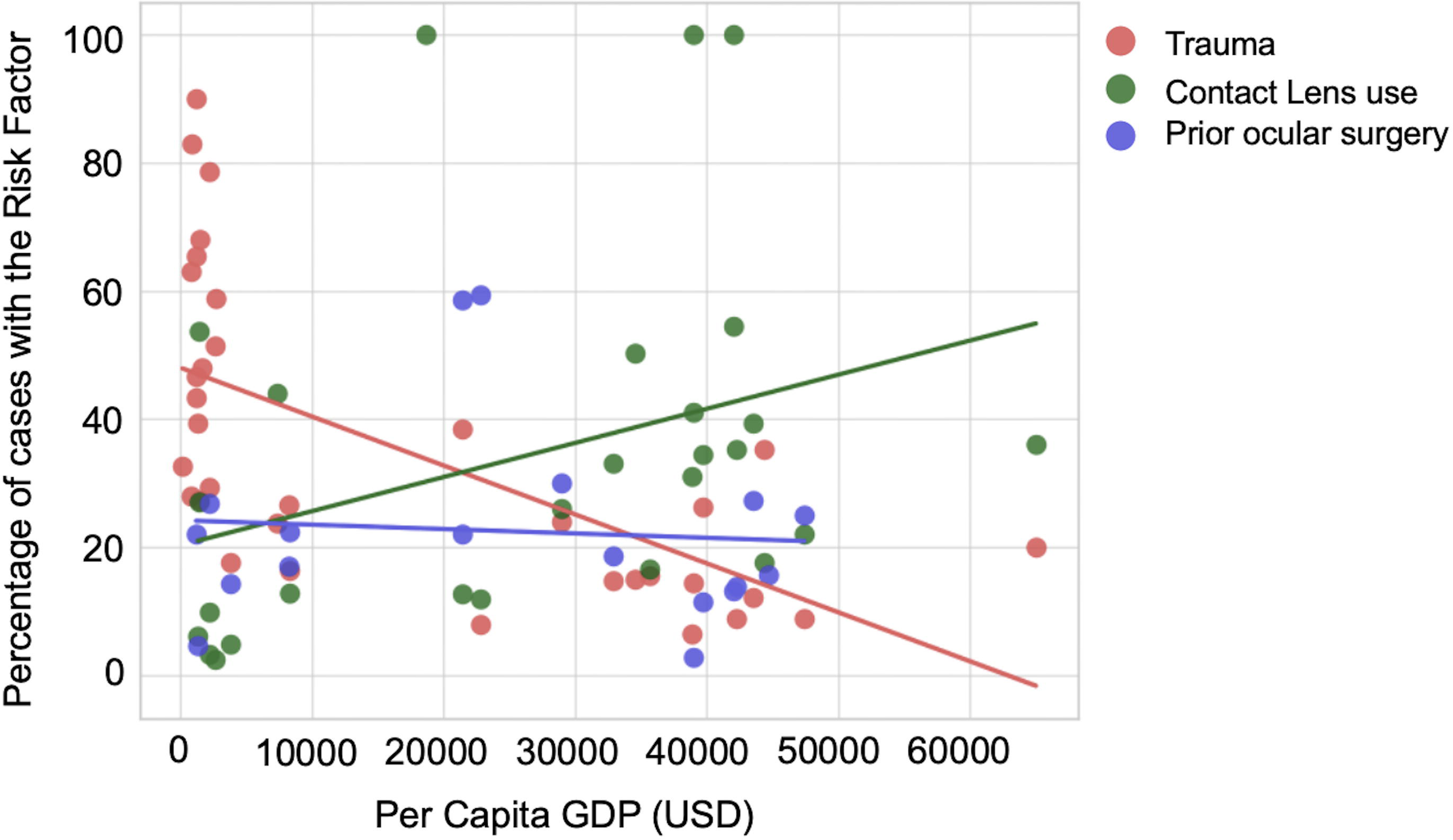
Scatterplot showing the relationship between national per capita GDP (USD) and the proportion of bacterial keratitis cases associated with three major risk factors: trauma (red), contact lens use (green), and prior ocular surgery (blue). Each dot represents data from an individual study. Linear trend lines indicate that trauma-related keratitis is more prevalent in lower-income settings, while contact lens–associated keratitis is increasingly common in higher-income countries. The proportion of cases related to prior ocular surgery remains relatively stable across GDP levels.

**Table 3.** Pooled prevalence of risk factors associated with bacterial keratitis from meta-analysis of included studies.

| Risk Factors | Case Numbers | Pooled Percentage |
| --- | --- | --- |
| Trauma | 10179 | 44.18% |
| Contact lens | 5233 | 22.71% |
| Prior ocular surgery | 3487 | 15.14% |
| Ocular surface disorder | 1844 | 8.00% |
| Dry eye | 604 | 2.62% |
| Trichiasis | 344 | 1.49% |
| Eroding suture | 291 | 1.26% |
| Diabetes Mellites | 279 | 1.21% |
| Postoperative SCL | 264 | 1.15% |
| Blepharitis | 226 | 0.98% |
| Previous keratitis | 183 | 0.80% |
| Trachoma | 80 | 0.35% |
| Herpes simplex | 15 | 0.06% |
| Glaucoma | 12 | 0.05% |

Among the 27 studies reporting antibiotic susceptibility (totaling ∼5,000 isolates), Gram-positive cocci demonstrated consistent susceptibility to vancomycin (>95% in nearly all studies), as well as high susceptibility to linezolid and teicoplanin (>98%). **(Figure 5 and Appendix S4)** Chloramphenicol was also effective against most Gram-positive organisms (∼92% susceptibility), while erythromycin and clindamycin showed moderate resistance rates; erythromycin susceptibility was only ∼57% in pooled data. For Gram-negative rods, aminoglycosides such as gentamicin, tobramycin, and amikacin showed excellent activity (>90% susceptibility), as did third- and fourth-generation cephalosporins like ceftazidime and cefepime. Carbapenems and piperacillin–tazobactam were effective in ≥85–90% of tested Gram-negative isolates.

**Figure 5.**
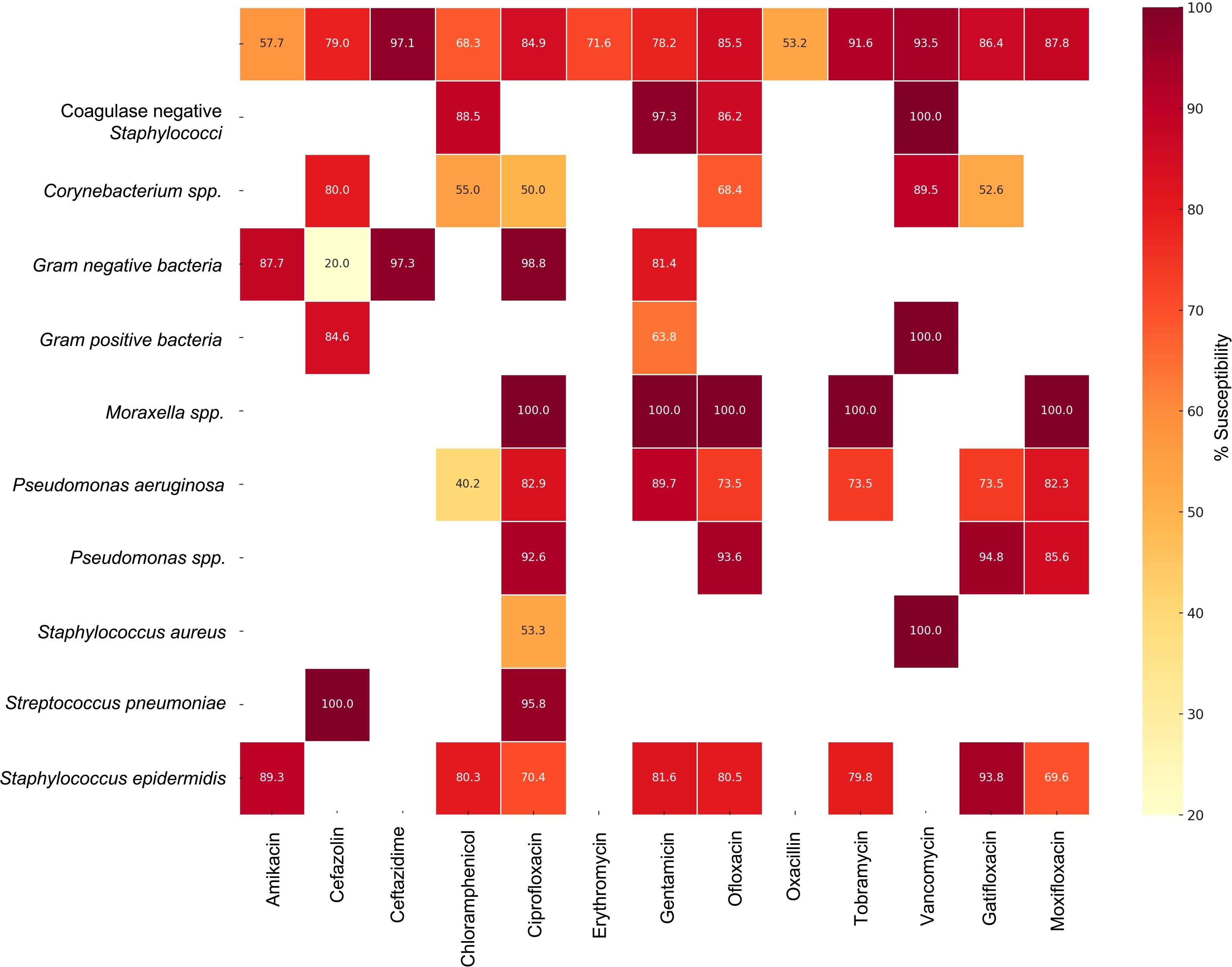
Heatmap summarizing pooled antibiotic susceptibility profiles of major bacterial pathogens isolated from infectious keratitis cases. Each cell shows the percentage of isolates susceptible to the corresponding antibiotic, color-coded from yellow (lower susceptibility) to dark red (higher susceptibility). Notable findings include high susceptibility of *Pseudomonas aeruginosa* to ceftazidime, gentamicin, and tobramycin; widespread resistance of *Staphylococcus aureus* to erythromycin; and excellent vancomycin susceptibility among gram-positive cocci.

Fluoroquinolone susceptibility was more variable and lower overall. Pooled ciprofloxacin susceptibility was 77% for *S. aureus* and 86–90% for *P. aeruginosa*, with more recent data showing downward trends. Levofloxacin had similar or slightly lower efficacy, while newer fluoroquinolones such as moxifloxacin and gatifloxacin had higher susceptibilities in the limited studies available (83% and 94%, respectively). These findings reflect growing resistance, especially among staphylococcal isolates. Methicillin resistance was also common: approximately 20% of *S. aureus* isolates were MRSA, with some centers reporting >30% MRSA prevalence. *Coagulase-negative Staphylococcus* isolates frequently demonstrated multidrug resistance, with up to 83% resistant to ≥3 antibiotic classes in certain series.

Overall, these susceptibility data suggest that empiric therapy for BK should include vancomycin for Gram-positive coverage and an aminoglycoside or third-generation cephalosporin for Gram-negative coverage, rather than relying solely on fluoroquinolones. Figure 3 presents a regional heatmap summarizing the percentage susceptibility of key antibiotic–pathogen combinations. Sensitivity analyses excluding lower-quality studies (NOS <4) did not significantly alter pooled BK prevalence estimates. The high heterogeneity observed across studies was expected given the global scope, variability in clinical settings, and differences in microbiological practices.

## Discussion

Our systematic review and meta-analysis provide a comprehensive global overview of BK, revealing important patterns in its prevalence, risk factors, and antimicrobial resistance. We found that approximately 44% of infectious keratitis cases globally are attributable to bacteria (pooled estimate), although the proportion ranged widely from ∼10% to 85% across studies. In temperate, high-income regions, BK constitutes most corneal infections. In North America an estimated 86–92% of corneal ulcers are bacterial, reflecting the relative rarity of fungal keratitis in cooler climates and settings with widespread contact lens use.^16–18^ In contrast, in tropical and low-income regions, fungal keratitis assumes a larger role. Our meta-analysis showed the lowest proportion of BK in Africa (∼29%), implying that non-bacterial etiologies (especially filamentous fungi) dominate in many African cases. This aligns with prior epidemiological observations that warm, humid climates foster fungal corneal infections (*Fusarium*, *Aspergillus*) associated with agricultural injuries.^15^ These findings reinforce the need for region-specific public health strategies. Notably, the World Health Organization has recognized the magnitude of corneal infections by proposing that infectious keratitis be designated a “Neglected Tropical Disease”, to promote awareness and allocate resources for prevention and treatment.^19^

Across >23,000 BK cases with reported risk factors, ocular trauma emerged as the most frequent precipitant (44% of cases), followed by contact lens wear (23%) and prior ocular surgery (15%). In Asia and Africa, corneal trauma, often from plant material or soil, is a ubiquitous trigger for keratitis.^20^ By contrast, in urban and high-income settings, contact lens use is a leading cause of corneal infection. We found a positive correlation between contact lens wear and BK, which is consistent with the well-documented risk of *Pseudomonas aeruginosa* infection in lens users.^21^ Prior studies have noted that in developed countries, contact lens wear accounts for as much as 30–65% of all infectious keratitis cases.^14,22^

Our analysis suggested an inverse correlation between a history of corneal surgery and bacterial keratitis. One interpretation is that post-surgical keratitis, when it occurs, may often involve atypical or opportunistic organisms, or that perioperative antibiotic prophylaxis protects against many bacterial infections.^23^ Additionally, some surgical cases, such as keratoplasty with suture infections might present with polymicrobial or fungal pathogens, lowering the proportion purely attributed to bacteria. Ocular surface disease (such as severe dry eye, exposure keratopathy) was another notable predisposing condition in several studies, particularly those from tertiary centers in higher-income countries.^24^ In a UK multi-center study the most common risk factors were ocular surface disorders (50% of cases) and contact lens wear (32%). Chronic surface diseases compromise corneal defenses and often lead to staphylococcal ulcers, as commensal flora invade the compromised epithelium.^23^ One of the most concerning findings of our review is the pattern of antibiotic susceptibility among keratitis pathogens, which points to emerging therapeutic challenges. While certain cornerstone antibiotics remain highly effective, others, notably the fluoroquinolones, show evidence of substantial resistance in ocular isolates.

Gram-positive isolates of the cornea remain uniformly susceptible to vancomycin in nearly all reported series. Our pooled data showed >95% susceptibility of staphylococci (including *S. aureus* and coagulase-negative strains) to vancomycin, with no reports of vancomycin resistance. Chloramphenicol also showed surprisingly high efficacy (∼90% susceptibility among Gram-positives in our review), in line with its broad spectrum against ocular commensals. This explains why chloramphenicol is still used empirically for mild keratitis in some regions (such as the UK), although it notably lacks activity against *P. aeruginosa*. By contrast, macrolides (such as erythromycin) and older-generation lincosamides had much lower efficacy (only ∼50–60% of staphylococci susceptible to erythromycin in pooled data), reflecting widespread resistance in those classes.

For Gram-negative organisms, our review found generally high susceptibility to traditional anti-pseudomonal antibiotics. Aminoglycosides such as gentamicin, tobramycin, and amikacin each showed >90% efficacy against *P. aeruginosa* and most Gram-negative rods in vitro. Likewise, third-generation cephalosporins (such as ceftazidime) and fourth-generation equivalents (cefepime) were highly active (often >90% susceptibility). Broad-spectrum penicillins with beta-lactamase inhibitors (like piperacillin–tazobactam) and carbapenems also retained strong activity (around 85–95% susceptibility in reported data). These findings suggest that the classic dual therapy for sight-threatening keratitis, combining a fortified aminoglycoside (for Gram-negatives) with a fortified cefazolin or vancomycin (for Gram-positives), remains a sound approach in the era of rising resistance.

Resistance trends in CoNS are also noteworthy. Several studies in our review reported high proportions of CoNS isolates resistant to multiple antibiotic classes. In some tertiary centers, over 80% of *S. epidermidis* keratitis isolates were non-susceptible to three or more classes of antibiotics (often including methicillin, fluoroquinolones, and macrolides). This is perhaps unsurprising given CoNS are common skin flora exposed to antibiotics and can easily exchange resistance genes. Although vancomycin covers these organisms, the rise of *multidrug-resistant CoNS* could pose problems in settings that lack access to fortified vancomycin or where chloramphenicol is used empirically (since many CoNS now produce chloramphenicol acetyltransferase enzymes conferring resistance). *Pseudomonas* resistance beyond fluoroquinolones appears less acute at present, most studies showed >95% of *P. aeruginosa* isolates were sensitive to tobramycin and ceftazidime. However, sporadic reports of increasingly virulent, drug-resistant pseudomonal strains (with efflux pump mutations or biofilm production) have emerged, so this remains a critical issue requiring continuous monitoring.^25,26^

Considering these findings, our meta-analysis underscores adopting a standardized approach to management. Empirical broad-spectrum coverage is justified for moderate to severe keratitis. Specifically, combining an anti-MRSA agent (like vancomycin) with an anti-pseudomonal agent (such as an aminoglycoside or third-generation cephalosporin) ensures coverage of the most prevalent and virulent bacteria in BK. Fluoroquinolone monotherapy should be used with caution. It may still be appropriate for small, peripheral ulcers or as initial outpatient therapy in low-risk cases, especially in settings where *MRSA prevalence is low*. But if there is no rapid improvement, escalation to fortified antibiotics or combination therapy is warranted. Lastly, surveillance of resistance patterns in ocular isolates needs to continue on a local and global scale.

## Strengths, Limitations, and Future Directions

Our study represents one of the largest collations of infectious keratitis data to date, spanning 58 studies from 32 countries and over 127,000 patients. By aggregating this evidence, we were able to derive globally relevant estimates that were previously lacking – for instance, the approximate 50:50 split between bacterial and non-bacterial keratitis worldwide, and the consistent top ranking of trauma and contact lens use as risk factors on different continents. We also synthesized antibiotic susceptibility from thousands of isolates, something no single study alone could accomplish, thereby providing a high-level map of resistance that can inform empiric treatment choices.

However, we acknowledge several limitations. Foremost, the heterogeneity between studies was high (I² ∼97% for pooled prevalence), reflecting diverse methodologies and populations. Our random-effects model accounted for some variation, but the summary estimates should be interpreted with caution given the variability. The studies spanned a long timeframe (1975–2022); changes in climate, healthcare access, and antibiotic usage over decades could influence the results. Older studies from the 1980s might report fewer contact lens infections (when fewer people wore lenses), whereas more recent studies may show higher resistance rates due to contemporary antibiotic pressures. We tried to mitigate this by subgroup analyses and sensitivity analyses (excluding lower-quality studies), which reassuringly did not drastically alter the pooled results, but some temporal trends may have been averaged out.

Another limitation is geographic sampling bias – certain high-burden regions had relatively few studies in our review (only 3 from Africa, for instance). This means our regional estimates (such as for Africa) have wider uncertainty and might not capture within-continent variation. It also highlights a research gap, as more population-based studies in Africa and rural Asia are needed, since our data relied mostly on tertiary-center case series that may underrepresent mild cases or those who never reached care. Relatedly, differences in healthcare infrastructure likely affected case ascertainment. In some low-resource settings, a large fraction of keratitis cases is managed empirically without lab confirmation; such cases might not have been counted as “bacterial” even if they were, simply due to lack of culture. Conversely, in specialized centers with advanced labs, even mild or atypical infections get cultured, potentially inflating the proportion of identified bacterial cases. We tried to include only studies with microbiological confirmation of diagnosis to ensure we were comparing similar outcomes (culture-proven etiology), but this criterion itself led to exclusion of some reports where culture data were incomplete. Thus, our 44% global BK prevalence should not be over-interpreted as a precise figure – rather, it is an estimate derived from the available literature. The true proportion of ulcers due to bacteria in the community could differ, especially where diagnostic capacity is limited.

We also did not perform a formal meta-analysis of antibiotic susceptibility rates due to methodological heterogeneity. The included studies used different antimicrobial panels, different breakpoints (some followed older CLSI vs newer EUCAST standards), and reported results in non-uniform ways (some by percentage, others by MIC ranges). Therefore, our statements on resistance are descriptive, summarizing general trends. While this yields useful insights, it lacks the precision of a quantitative meta-analysis. Clinicians should interpret the antibiotic efficacy categories (high ≥90%, moderate 50–89%, low <50% susceptible) in the context of their local formulary and resistance data. A related point is that susceptibility does not equal clinical outcome in vivo, factors like drug penetration, biofilm formation, and pathogen virulence mean that even a “susceptible” infection can sometimes fail to respond to treatment. We did not have patient-level data to analyze clinical cure rates by antibiotic, which would be an excellent area for future research.

Despite these limitations, our review provides a needed global perspective on bacterial keratitis. The findings emphasize that BK is a significant contributor to corneal blindness worldwide, but its epidemiology varies widely by region and context. Going forward, a few key areas deserve attention: 1) Improved surveillance and reporting: There is a need for standardized reporting of infectious keratitis causes and outcomes, especially from underrepresented regions. National registries or multicenter studies are valuable models. 2) Preventive interventions: Given that a large share of BK is associated with modifiable risk factors, targeted prevention could yield major public health gains. Simple measures like farmer education on eye protection, enforcement of workplace safety goggles, and public awareness campaigns on contact lens hygiene could reduce the incidence of keratitis.

In conclusion, this global review of bacterial keratitis highlights that while the burden of BK is immense, it is not monolithic, geographic, and climatic factors shape its prevalence and etiological agents. The data on antimicrobial resistance, especially the erosion of fluoroquinolone efficacy and persistence of MRSA, suggest that the era of easy, one-size-fits-all treatment is waning. Tailored, context-aware management, choosing therapy based on local epidemiology and risk factor profile, is the way forward to optimize outcomes in bacterial keratitis.

## Data Availability

All data produced in the present study are available upon reasonable request to the authors

