## Appendix S1 for "Global Burden, Risk Factors, Causative Organisms and Antibiotic Susceptibility Patterns in Bacterial Keratitis"

**Appendix S1: SEARCH STRATEGY**

We will conduct a comprehensive electronic search across the following databases: PubMed (1974 to July 1, 2022), PubMed Central, SCOPUS, and APASyncINFO including Epub Ahead of Print, In-Process & Other Non-Indexed Citations, Daily, and Versions® (1974 to July 1, 2022). The search will be conducted using Ovid’s interface and include both controlled vocabulary (MeSH and Emtree terms) and free-text keywords.

1. bacterial keratitis [Including Related Terms]

2. microbial keratitis [Including Related Terms]

3. infectious keratitis [Including Related Terms]

4. suppurative keratitis [Including Related Terms]

5. corneal ulcer [Including Related Terms]

6. bacterial corneal ulcer [Including Related Terms]

7. corneal infection [Including Related Terms]

8. Gram-positive bacteria AND keratitis

9. Gram-negative bacteria AND keratitis

10. 1 OR 2 OR 3 OR 4 OR 5 OR 6 OR 7 OR 8 OR 9

11. prevalence [Including Related Terms]

12. incidence [Including Related Terms]

13. burden [Including Related Terms]

14. epidemiology [Including Related Terms]

15. rate [Including Related Terms]

16. risk factor* [Including Related Terms]

17. antimicrobial susceptibility OR antibiotic resistance

18. 11 OR 12 OR 13 OR 14 OR 15 OR 16 OR 17

19. 10 AND 18

Searches were not limited by geographic location or publication year but will be restricted to articles published in English. The reference lists of included studies and relevant systematic reviews were manually screened to identify additional eligible studies.
