## Appendix S2 for "Global Burden, Risk Factors, Causative Organisms and Antibiotic Susceptibility Patterns in Bacterial Keratitis"

**Appendix S2:** Quality assessment of included studies using an adapted Grading of Recommendations Assessment, Development and Evaluation (GRADE) framework for prevalence studies

| Author and Year | Diagnostic accuracy (/2) | Patient n>30 (/1) | Up to date (/2) | %BK/MK (/1) | Overall score (/6) |
| --- | --- | --- | --- | --- | --- |
| (Abdullatif Al-Raisi, 2018) | 1 | 1 | 2 | 1 | 5 |
| (Alexandrakis, 2000) | 1 | 0 | 0 | 1 | 2 |
| (Al-Hazzaa & Tabbara, 1988) | 1 | 1 | 0 | 1 | 3 |
| (Almizel et al., 2019) | 1 | 1 | 2 | 1 | 5 |
| (Al-Shakarchi, 2007) | 1 | 1 | 0 | 1 | 3 |
| (Al-Shehri et al., 2008) | 1 | 1 | 0 | 1 | 3 |
| (Asbell, 1982) | 1 | 1 | 0 | 1 | 3 |
| (Badawi et al., 2017) | 1 | 1 | 2 | 1 | 5 |
| (Bamdad et al., 2015) | 1 | 1 | 1 | 1 | 4 |
| (Basak et al., 2005) | 1 | 1 | 0 | 1 | 3 |
| (Basak et al., 2005) | 1 | 1 | 0 | 1 | 3 |
| (Bharathi et al., 2007) | 1 | 1 | 0 | 1 | 3 |
| (Bharathi et al., 2009) | 1 | 1 | 0 | 1 | 3 |
| (Bourcier, 2003) | 1 | 1 | 0 | 1 | 3 |
| (Cabrera‐Aguas et al., 2019) | 2 | 1 | 2 | 1 | 6 |
| (Cariello et al., 2011) | 1 | 1 | 2 | 1 | 5 |
| (Chen, 2008) | 1 | 1 | 0 | 1 | 3 |
| (Chidambaram et al., 2018) | 1 | 1 | 2 | 1 | 5 |
| (Das et al., 2015) | 1 | 0 | 1 | 1 | 3 |
| (Das et al., 2019) | 1 | 1 | 2 | 1 | 5 |
| (Das, 2006) | 1 | 1 | 0 | 1 | 3 |
| (Deorukhkarl et al., 2012) | 2 | 1 | 1 | 1 | 5 |
| (Fong et al., 2004) | 2 | 1 | 0 | 1 | 4 |
| (Fong et al., 2007) | 1 | 1 | 0 | 1 | 3 |
| (Galentine et al., 1984) | 1 | 1 | 0 | 1 | 3 |
| (Goldstein et al., 1999) | 1 | 1 | 0 | 1 | 3 |
| (Hernandez-Camarena et al., 2015) | 1 | 1 | 1 | 1 | 4 |
| (Hooi, 2005) | 1 | 1 | 0 | 1 | 3 |
| (Hsiao et al., 2016) | 1 | 1 | 1 | 1 | 4 |
| (Ibrahim et al., 2009) | 1 | 1 | 0 | 1 | 3 |
| (Inoue et al., 2007) | 1 | 1 | 0 | 1 | 3 |
| (Jin et al., 2017) | 1 | 1 | 2 | 1 | 5 |
| (Kaliamurthy et al., 2013) | 1 | 1 | 1 | 1 | 4 |
| (Kaye et al., 2013) | 1 | 1 | 1 | 1 | 4 |
| (Khor et al., 2018) | 1 | 1 | 2 | 1 | 5 |
| (Lalitha et al., 2014) | 1 | 1 | 1 | 1 | 4 |
| (Lalitha et al., 2016) | 1 | 1 | 1 | 1 | 4 |
| (Lam et al., 2002) | 2 | 1 | 0 | 1 | 4 |
| (Laspina et al., 2004) | 1 | 1 | 0 | 1 | 3 |
| (Lichtinger et al., 2012) | 1 | 1 | 1 | 1 | 4 |
| (Lin et al., 2012) | 1 | 1 | 1 | 1 | 4 |
| (Lin et al., 2016) | 1 | 1 | 1 | 1 | 4 |
| (Marasini et al., 2016) | 1 | 1 | 1 | 1 | 4 |
| (Mun et al., 2019) | 1 | 1 | 2 | 1 | 5 |
| (Ni et al., 2015) | 1 | 1 | 1 | 1 | 4 |
| (Orlans et al., 2011) | 1 | 1 | 0 | 1 | 3 |
| (Pan et al., 2016) | 2 | 1 | 1 | 1 | 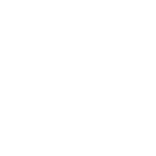<https://www.tandfonline.com/doi/full/10.1080/09286586.2018.1454964>5 |
| (Pandita & Murphy, 2011) | 2 | 1 | 0 | 1 | 4 |
| (Peng et al., 2018) | 1 | 1 | 2 | 1 | 5 |
| (Politis et al., 2016) | 1 | 1 | 1 | 1 | 4 |
| (Rautaraya et al., 2014) | 1 | 1 | 1 | 1 | 4 |
| (Roth et al., 2022) | 1 | 1 | 2 | 1 | 5 |
| (Saillard et al., 2018) | 2 | 1 | 2 | 1 | 6 |
| (Schaefer, 2001) | 2 | 1 | 0 | 1 | 4 |
| (Seal et al., 1999) | 1 | 1 | 0 | 1 | 3 |
| (Shalchi et al., 2011) | 1 | 1 | 0 | 1 | 3 |
| (Sharma et al., 2003) | 1 | 0 | 0 | 1 | 2 |
| (Sharma et al., 2009) | 1 | 1 | 0 | 1 | 3 |
| (Song et al., 2012) | 2 | 1 | 1 | 1 | 5 |
| (Sood et al., 2012) | 1 | 1 | 1 | 1 | 4 |
| (Srinivasan et al., 1997) | 2 | 1 | 0 | 1 | 4 |
| (Sun, 2004) | 1 | 1 | 0 | 1 | 3 |
| (Sun, 2004) | 1 | 1 | 0 | 1 | 3 |
| (Tabbara, 2000) | 1 | 0 | 0 | 1 | 2 |
| (Tam et al., 2017) | 1 | 1 | 2 | 1 | 5 |
| (Tan et al., 2017) | 1 | 1 | 2 | 1 | 5 |
| (Termote et al., 2018) | 2 | 1 | 2 | 1 | 6 |
| (Ting et al., 2018) | 1 | 1 | 2 | 1 | 5 |
| (Ting et al., 2021) | 1 | 1 | 2 | 1 | 5 |
| (Tobimatsu et al., 2018) | 1 | 0 | 2 | 1 | 4 |
| (Toshida et al., 2007) | 1 | 1 | 0 | 1 | 3 |
| (Tuft, 2000) | 1 | 1 | 0 | 1 | 3 |
| (Upadhyay et al., 1991) | 1 | 1 | 0 | 1 | 3 |
| (Verhelst et al., 2006) | 1 | 1 | 0 | 1 | 3 |
| (Wang et al., 1998) | 1 | 1 | 0 | 1 | 3 |
| (Wong, 2003) | 1 | 1 | 0 | 1 | 3 |
| (Yeh et al., 2006) | 1 | 1 | 0 | 1 | 3 |
| (Yilmaz et al., 2007) | 1 | 1 | 0 | 1 | 3 |
