## Appendix S3 for "Global Burden, Risk Factors, Causative Organisms and Antibiotic Susceptibility Patterns in Bacterial Keratitis"

**Appendix S3:** Reported Prevalence of Bacterial Isolates in Bacterial Keratitis Across Included Studies, Categorized by Gram Classification and Species

|  | Gram  Negative  Bacilli | | | | | | | Gram Negative Cocci | Gram  Positive  Bacilli | | | Gram  Positive  Cocci | | | | | | | |
| --- | --- | --- | --- | --- | --- | --- | --- | --- | --- | --- | --- | --- | --- | --- | --- | --- | --- | --- | --- |
|  | Acinetobacter baumanii | Enterobactor spp | Hemophilus influenzae | Klebsiella oxytoca | Proteus mirabilis | Pseudomonas aeruginosa | Serratia marcescens | Moraxella spp | Corynebacterium spp. | Nocardia spp. | Miccrococcus spp. | | Staphylococcus aureus | Staphylococcus epidermidis | Staphylococcus homini | Staphylococcus simulans | Stretococcus pneumoniae | Streptococcus pyogenes | Streptococcus viridans |
| (Abdullatif Al-Raisi, 2018) |  |  |  |  |  | 54 |  |  |  |  |  | |  | 29 |  |  | 47 |  | 13 |
| (Alexandrakis, 2000) |  |  |  |  |  | 25.7 | 7.6 |  |  |  |  | | 19.4 |  |  |  |  |  |  |
| (Al-Hazzaa & Tabbara, 1988) |  |  | 4 |  |  | 12 |  | 4 |  |  |  | | 4 | 21 |  |  | 26 |  |  |
| (Al-Shakarchi, 2007) |  |  |  |  |  | 42 |  | 1.2 |  |  |  | | 19.1 | 24.1 |  |  | 9.3 |  |  |
| (Al-Shehri et al., 2008) |  |  |  |  |  | 28 |  | 16 |  |  |  | | 12 | 56 |  |  | 31 |  |  |
| (Asbell, 1982) |  |  |  |  |  | 8 |  | 16 |  |  |  | | 48 |  |  |  | 8 |  |  |
| (Badawi et al., 2017) |  |  |  |  |  | 9 |  | 1 |  |  |  | | 18 | 6 |  |  | 3 |  |  |
| (Basak et al., 2005) |  | 2.6 |  |  |  | 21.1 |  | 1.6 |  |  |  | | 42.6 | 15.7 |  |  | 9.4 |  |  |
| (Bharathi et al., 2007) |  |  |  |  |  | 17.3 |  | 9 |  |  | 6 | | 4.3 | 21.6 |  |  | 35.95 |  |  |
| (Bharathi et al., 2009) |  |  |  |  |  | 19.7 |  | 0.7 |  |  |  | | 3.8 | 18.3 |  |  | 36 |  |  |
| (Cariello et al., 2011) |  |  |  |  |  | 11.77 |  | 4.74 |  |  |  | | 21.1 | 26.3 |  |  | 7.5 |  |  |
| (Chidambaram et al., 2018) |  |  |  |  |  | 21 |  |  |  | 16 |  | |  |  |  |  | 47 |  |  |
| (Das et al., 2019) |  |  |  |  |  | 45.2 |  |  |  |  |  | | 16.4 |  |  |  | 19.3 |  |  |
| (Deorukhkarl et al., 2012) |  |  |  |  |  | 7.96 |  | 3.98 |  |  |  | | 17.25 | 10.61 |  |  | 32.74 |  |  |
| (Fong et al., 2004) |  |  |  |  |  | 33.7 |  |  |  |  |  | | 8.4 |  |  |  |  |  |  |
| (Galentine et al., 1984) |  |  |  |  |  | 23 |  |  |  |  |  | | 12.5 | 7.14 |  |  | 1.78 |  |  |
| (Gopinathan et al., India 2009) |  |  | 1 |  |  | 9.7 |  | 1.4 | 14.5 | 1.8 |  | | 5.3 | 32.5 |  |  | 13.9 | 6.5 |  |
| (Hernandez-Camarena et al., 2015) |  |  |  |  |  | 12 |  | 1 |  |  |  | | 9 | 25 |  |  |  |  |  |
| (Hsiao et al., 2016) |  |  |  |  |  | 24.4 | 5.2 |  |  |  |  | | 8.4 | 16.6 |  |  | 2.9 |  |  |
| (Ibrahim et al., 2009) |  |  |  |  |  | 12 |  |  |  |  |  | | 11.5 | 31.7 |  |  | 4.7 |  |  |
| (Jin et al., 2017) |  |  |  |  |  | 19.8 |  |  |  |  |  | | 3.1 | 15.6 |  |  | 6.3 |  |  |
| (Kaliamurthy et al., 2013) |  |  |  |  |  |  |  |  |  |  |  | | 19.5 | 44 |  |  | 11.6 |  |  |
| (Khor et al., 2018) |  |  |  |  |  | 10.7 |  |  |  |  |  | |  |  |  |  | 6.3 |  |  |
| (Lalitha et al., 2014) |  |  |  |  |  | 5.4 |  |  |  |  |  | | 1.2 |  |  |  | 7 |  |  |
| (Lalitha et al., India 2016) |  |  |  |  |  | 24.3 |  |  | 6.4 | 6.7 |  | | 5.3 | 4.1 |  |  | 32.7 | 5.8 | 5.8 |
| (Lam et al., 2002) |  |  |  |  |  | 31.3 |  |  |  |  |  | | 10 |  |  |  |  | 3.33 |  |
| (Laspina et al., 2004) |  |  |  |  |  | 10.7 |  |  |  |  |  | | 23.72 | 25.2 |  |  |  |  |  |
| (Lichtinger et al., 2012) |  |  |  |  |  | 10 |  | 5 |  |  |  | | 17 | 37 |  |  |  |  |  |
| (Lin et al., 2012) |  |  |  |  |  | 24.3 |  |  |  |  |  | |  |  |  |  | 35.1 |  |  |
| (Lin et al., 2016) |  |  |  |  |  | 12.4 |  |  |  |  |  | |  | 31.9 |  | 5.53 |  |  |  |
| (Mun et al., 2019) |  |  |  |  |  | 10.3 |  |  | 5.6 |  |  | | 12.1 | 15.9 |  |  | 8.4 |  | 5.6 |
| (Ni et al., 2015) |  |  |  |  |  | 9.29 |  |  |  |  |  | | 8.9 | 6.5 |  |  |  |  |  |
| (Orlans et al., 2011) |  |  |  |  |  | 24.3 |  | 2.6 | 6 |  |  | | 14.3 | 25.8 |  |  | 3 |  |  |
| (Pan et al., 2016) |  |  |  |  |  | 42.86 |  |  |  |  |  | | 48.57 |  |  |  |  |  |  |
| (Pandita & Murphy, 2011) |  |  |  |  |  | 3.4 |  | 8 |  |  |  | | 11.5 | 40.8 |  |  | 7.5 |  |  |
| 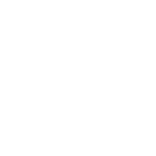<https://www.tandfonline.com/doi/full/10.1080/09286586.2018.1454964>  (Rautaraya et al., 2014) |  |  |  |  |  | 8.3 |  |  |  |  |  | | 18.4 |  |  |  | 24.7 |  |  |
| (Roth et al., 2022) |  |  |  |  |  | 18.1 |  |  |  |  |  | | 16 | 14.1 |  |  | 4.6 |  |  |
| (Shalchi et al., 2011) |  |  |  |  |  | 49.4 |  |  |  |  |  | | 14.8 | 8 |  |  |  |  |  |
| (Sharma et al., 2003) |  |  |  |  |  | 35.7 |  |  |  |  |  | |  | 14.3 |  |  | 7.1 |  |  |
| (Sharma et al., 2009) |  |  |  |  |  | 9.7 |  | 1.4 |  |  |  | | 5.3 | 32.5 |  |  | 13.9 |  |  |
| (Song et al., 2012) |  |  |  |  |  | 2.6 |  |  |  |  |  | | 2.6 | 35.9 |  |  |  |  |  |
| (Srinivasan et al., 1997) |  |  |  |  |  | 14.4 |  | 1.8 | 12.5 |  |  | |  | 10.2 |  |  | 44.3 |  |  |
| (Sun et al, 2004) | 1.4 |  |  |  |  | 32.2 |  | 0.2 |  |  | 11 | | 5.5 | 18.6 |  |  |  |  |  |
| (Tam et al., 2017) |  |  |  |  |  | 10 |  | 5 |  |  |  | | 15 | 37 |  |  | 15 |  |  |
| (Tan et al., 2017) |  |  |  |  |  | 37.1 |  | 22.1 |  |  |  | | 23.9 | 38.5 |  |  |  |  |  |
| (Termote et al., 2018) |  |  |  |  |  | 5.15 |  | 6.75 |  |  |  | | 15.47 | 28.57 |  |  |  |  |  |
| (Tewari et al., 2012) |  |  |  |  |  | 18.9 |  |  |  |  |  | | 32.7 | 25.8 |  |  |  |  |  |
| (Toshida et al., 2007) |  |  |  |  |  | 1 | 7.8 |  |  |  |  | | 13.7 | 28.4 |  |  |  |  |  |
| (Upadhyay et al., 1991) |  |  |  |  |  | 10.6 |  | 2 |  |  |  | | 10.8 | 11.5 |  |  | 31.1 |  |  |
| (Verhelst et al., 2006) |  |  |  | 9.9 |  | 53.46 | 15.8 |  |  |  |  | | 8.9 |  |  |  |  |  |  |
| (Wong et al., 2003) |  |  |  |  |  | 5.73 |  | 4.92 |  |  |  | |  | 11.48 |  |  | 9.02 |  |  |
| (Yilmaz et al., 2007) |  | 2.2 |  |  |  | 6.6 |  |  |  |  |  | | 24.4 | 26.6 |  |  | 15.55 |  |  |
| (Zhang et al., China 2008) |  |  |  |  | 0.72 | 20.07 | 1.43 |  | 16.85 |  | 9.68 | | 7.17 | 13.98 |  |  | 7.53 | 2.15 |  |
