## Appendix S4 for "Global Burden, Risk Factors, Causative Organisms and Antibiotic Susceptibility Patterns in Bacterial Keratitis"

**Appendix S4:** Antibiotic susceptibility patterns of bacterial isolates from keratitis cases reported across included studies.

| Bacterial | Study | Antibiotic | | | | | | | | | | | | |
| --- | --- | --- | --- | --- | --- | --- | --- | --- | --- | --- | --- | --- | --- | --- |
|  |  | Amikacin | Cefazolin | Ceftazidime | Chloramphenicol | Ciprofloxacin | Erythromycin | Gatifloxacin | Gentamicin | Moxifloxacin | Ofloxocin | Oxacillin | Tobramycin | Vancomycin |
| Corynebacterium spp | Das et al., 2015) |  | 80.0% |  | 55.0% | 50.0% |  | 52.6% |  |  | 68.4% |  |  | 89.5% |
|  | Alfonso et al., 1986) |  |  |  | 3.0% | 1.0% |  |  | 0.0% |  |  |  |  |  |
|  | Jin et al., 2017 |  |  |  |  |  | 67% |  |  |  |  |  | 87% | 100% |
| Coagulase-Negative Staphylococci | (Tan et al., 2017) |  |  |  | 88.5% |  |  |  | 97.3% |  | 86.2% |  |  | 100.0% |
|  | Termote et al., 2018) |  |  |  |  | 65.0% |  |  | 84.0% | 81.0% | 65.0% |  | 85.0% |  |
|  | Lichtinger et al., 2012) |  | 46.7% |  |  |  | 49.0% |  |  |  |  | 56.9% |  | 100.0% |
|  | Hernandez-Camarena et al., 2015) |  | 88.0% |  |  | 95.0% |  |  | 92.0% |  | 93.0% | 44.0% |  |  |
|  | (Ni et al., 2015) |  |  |  |  |  |  | 68.0% |  | 68.0% |  |  |  |  |
|  | (Jin et al., 2017) |  |  |  |  |  |  |  |  |  |  |  |  |  |
|  | (Lalitha et al., 2016) |  |  |  |  |  |  |  | 48.3% |  | 43.3% |  |  |  |
|  | Alfonso et al., 1986) |  |  |  | 16.0% | 37.0% |  |  | 21.0% |  |  |  |  |  |
| S. pneumoniae | Das et al., 2019) |  | 100.0% |  |  | 95.8% |  |  |  |  |  |  |  |  |
|  | Kaliamurthy et al., 2013) | 37.8% |  |  | 82.1% | 75.7% |  | 95.0% | 30.7% | 86.5% | 86.4% |  | 42.8% |  |
|  | Lalitha et al., 2016) |  |  |  | 96.9% |  |  |  |  |  | 95.5% |  |  | 100.0% |
|  | (Peng et al., 2018) |  |  |  |  |  | 76.0% |  |  | 100.0% | 100.0% |  |  | 100.0% |
|  | Jin et al., 2017 |  |  |  |  |  | 67% |  |  |  |  |  |  |  |
| S. aureus | Das et al., 2019) |  |  |  |  | 53.3% |  |  |  |  |  |  |  | 100.0% |
|  | Kaliamurthy et al., 2013) | 95.3% |  |  | 84.7% | 76.6% |  | 97.9% | 84.7% | 73.4% | 83.0% |  | 86.4% |  |
|  | Saillard et al., 2018) |  |  | 100.0% |  |  |  |  |  |  |  |  |  | 100.0% |
|  | (Tan et al., 2017) |  |  |  | 95.6% |  |  |  | 99.6% |  | 91.2% |  |  | 100.0% |
|  | Galentine et al., 1984) |  |  |  |  | 100.0% |  |  |  |  | 87.3% |  |  |  |
|  | Lichtinger et al., 2012) |  | 98.1% |  |  |  |  |  |  |  |  | 98.7% |  | 100.0% |
|  | Yeh et al., 2006) |  | 76.0% |  |  |  |  |  |  |  |  |  |  | 100.0% |
|  | Hernandez-Camarena et al., 2015) |  | 79.0% |  |  | 89.0% |  |  | 72.0% |  | 89.0% | 21.0% |  |  |
|  | Jin et al., 2017 |  |  |  |  |  | 67% |  |  |  |  |  |  |  |
| Pseudomonas spp. | Das et al., 2019) |  |  |  |  | 92.6% |  | 94.8% |  | 85.6% | 93.6% |  |  |  |
|  | (Peng et al., 2018) |  |  | 98.0% |  |  |  |  | 94.0% |  |  |  | 94.0% |  |
|  | Yeh et al., 2006) |  |  | 95.0% |  | 94.8% |  |  | 97.5% |  |  |  |  |  |
|  | (Tan et al., 2017) |  |  | 98.7% |  | 100.0% |  |  | 100.0% |  | 95.0% |  | 100.0% |  |
| Pseudomonas aeruginosa | Kaliamurthy et al., 2013) | 89/7% |  |  | 40.2% | 82.9% |  | 73.5% | 89.7% | 82.3% | 73.5% |  | 73.5% |  |
|  | Lalitha et al., 2016) |  |  | 94.5% |  | 86.9% |  |  |  |  | 86.9% |  |  |  |
|  | Saillard et al., 2018) | 100.0% |  | 100.0% |  |  |  |  |  |  |  |  |  |  |
|  | Galentine et al., 1984) |  |  |  |  | 100.0% |  |  |  |  | 98.2% |  |  |  |
|  | (Lichtinger et al., 2012) |  |  |  |  | 97.8% |  |  | 98.9% |  |  |  |  |  |
|  | (Hernandez-Camarena et al., 2015) |  |  |  |  | 100.0% |  |  | 88.0% |  | 86.0% | 25.0% |  |  |
|  | (Ni et al., 2015) |  |  |  |  | 100.0% |  | 88.0% |  | 96.0% |  |  |  |  |
| Staphylococcus epidermidis | Kaliamurthy et al., 2013) | 89.3% |  |  | 80.3% | 70.4% |  | 93.8% | 81.6% | 69.6% | 80.5% |  | 79.8% |  |
|  | Saillard et al., 2018) |  |  |  |  |  |  |  |  |  |  |  | 100.0% |  |
|  | Hernandez-Camarena et al., 2015) |  | 74.0% |  |  | 61.0% |  |  | 50.0% |  | 62.0% | 56.0% |  |  |
| Moraxella species | Termote et al., 2018) |  |  |  |  | 100.0% |  |  | 100.0% | 100.0% | 100.0% |  | 100.0% |  |
|  | (Das, 2006) |  |  |  |  |  |  | 100% |  | 100% |  |  |  |  |
| Gram-Negative Bacteria | Fong et al., 2007) | 87.7% | 20.0% | 97.3% |  | 98.8% |  |  | 81.4% |  |  |  |  |  |
|  | (Marasini et al., 2016) |  | 100.0% | 100.0% | 94.7% | 100.0% | 100.0% |  | 100.0% |  |  |  | 100.0% | 0.0% |
|  | (Mun et al., 2019) | 0.0% |  |  | 87.0% |  |  |  | 75.0% | 91.7% |  |  |  |  |
|  | (Orlans et al., 2011) |  |  | 100.0% | 38.6% | 99.0% |  |  | 100.0% |  |  |  |  |  |
|  | Saillard et al., 2018) | 0.0% |  | 100.0% |  |  |  |  |  |  |  |  |  | 100.0% |
|  | Schaefer, 2001) |  |  |  |  |  |  |  |  |  |  |  |  |  |
|  | Tam et al., 2017) |  |  | 91.4% |  | 96.1% |  |  | 95.6% |  |  |  | 93.5% |  |
|  | (Tan et al., 2017) | 97.9% |  | 92.1% | 75.6% | 98.9% |  |  | 96.5% |  | 90.8% |  |  |  |
|  | Termote et al., 2018) |  |  |  | 50.0% |  |  |  |  |  |  |  |  |  |
|  | Galentine et al., 1984) |  |  |  |  | 96.3% |  |  |  |  | 96.3% |  |  |  |
|  | Hsiao et al., 2016) | 88.3% |  | 90.9% |  | 93.7% |  |  | 84.7% |  |  |  |  |  |
|  | (Pandita & Murphy, 2011) |  |  |  | 98.3% |  |  |  |  |  |  |  | 100.0% |  |
|  | (Rautaraya et al., 2014) | 100.0% |  |  |  | 90.0% |  | 95.1% |  |  |  |  |  |  |
|  | Yeh et al., 2006) |  |  |  |  | 100.0% |  |  | 93.0% |  |  |  |  |  |
|  | Lichtinger et al., 2012) |  |  | 98.2% |  | 97.4% |  |  | 96.8% |  |  |  | 98.2% |  |
|  | Alfonso et al., 1986) |  |  |  | 3.0% | 68.0% |  | 61.0% |  |  |  |  |  |  |
|  | (Shalchi et al., 2011) |  |  |  | 25.9% |  |  |  |  |  |  |  |  |  |
| Gram-Positive Bacteria | Fong et al., 2007) |  | 84.6% |  |  |  |  |  | 63.8% |  |  |  |  | 100.0% |
|  | (Marasini et al., 2016) |  | 100.0% | 100.0% | 100.0% | 98.8% | 100.0% |  | 100.0% |  |  |  | 100.0% |  |
|  | (Mun et al., 2019) | 0.0% | 53.8% |  | 86.0% |  |  |  | 69.7% | 93.8% |  |  | 100.0% | 100.0% |
|  | (Orlans et al., 2011) |  |  |  | 92.7% | 85.4% |  |  | 87.2% |  |  |  |  | 100.0% |
|  | Schaefer, 2001) |  |  |  |  |  |  |  |  |  |  |  |  |  |
|  | Tam et al., 2017) |  | 65.8% |  |  |  | 55.5% |  |  |  |  |  |  | 99.6% |
|  | (Tan et al., 2017) |  |  |  | 92.1% |  |  |  | 88.8% |  | 83.1% |  |  | 99.8% |
|  | Tuft, 2000) |  |  |  |  |  |  |  |  |  | 96.8% |  |  |  |
|  | Galentine et al., 1984) |  |  |  |  | 77.5% |  |  |  |  | 81.9% |  |  |  |
|  | (Lichtinger et al., 2012) |  | 70.4% |  |  |  | 62.7% |  |  |  |  | 70.9% |  | 100.0% |
|  | (Pandita & Murphy, 2011) |  |  |  | 95.5% |  |  |  |  |  |  |  | 95.5% |  |
|  | (Rautaraya et al., 2014) |  | 96.2% |  | 88.9% |  |  | 86.5% |  |  |  |  |  | 96.5% |
|  | Alfonso et al., 1986) |  |  |  | 28.0% | 65.0% |  |  | 41.0% |  |  |  |  |  |
